# Intrathrombus NETosis Defines Prognostically Distinct Phenotypes in Large-Vessel Occlusion Stroke

**DOI:** 10.64898/2026.10.04.26364710

**Authors:** B Díaz-Benito, P Calleja, F Ostos, A Ruiz-García, L Alzamora, A Martínez-Salio, M Gutiérrez-Sánchez, F Ballenilla, J Álvarez-Cienfuegos, C Gubern, Y Silva, A García-Culebras, MA Moro, A Moraga, I Lizasoain

## Abstract

**Background:** Immunothrombosis is increasingly recognized in large-vessel-occlusion acute ischemic stroke (LVO-AIS), yet most biomarker studies infer thrombus biology from circulating neutrophil extracellular trap (NET) markers or from pre-defined thrombus subtypes. We prospectively studied 160 patients with anterior-circulation LVO-AIS treated with mechanical thrombectomy to determine whether intrathrombus NETosis markers identify thrombus phenotypes with distinct clinical trajectories, and whether plasma NETosis biomarkers reflect the molecular composition of thrombi.

**Methods:** Thrombi underwent quantitative histopathology and immunohistochemistry for citrullinated histone H3 (CitH3), neutrophil elastase (NE) and CD61. Thrombi were analyzed without prior etiological or histological stratification, and intrathrombus markers were dichotomized at cohort medians (CitH3 6.62%, NE 3.95%, CD61 23.09%) to define internally standardized candidate phenotypes. Associations with thrombus composition, collateral circulation (Tan 2-3), 3-month functional independence (modified Rankin Scale 2 or less) and long-term mortality were assessed by correlation analyses and progressively adjusted multivariable models, with cross-compartment correlations corrected by the Benjamini-Hochberg false discovery rate (FDR).

**Results:** Intrathrombus markers defined prognostically distinct phenotypes: CD61 tracked functional recovery, NE preserved collateral circulation and CitH3 long-term survival, each remaining significant after multivariable adjustment. Circulating NE, myeloperoxidase and leukocyte count instead tracked the macroscopic red-blood-cell-fibrin composition of retrieved thrombi. Cross-compartment concordance was limited: after FDR correction the only significant plasma-thrombus association was an inverse correlation between plasma cell-free DNA and intrathrombus NE, and no intrathrombus marker correlated positively with its circulating counterpart.

**Conclusions:** Local immunothrombotic signatures therefore provide clinically relevant information not captured by plasma markers, and median-defined intrathrombus thresholds represent hypothesis-generating cutoffs for external validation and NET-targeted treatment stratification.

## Introduction

Acute ischemic stroke thrombi are highly heterogeneous structures whose composition influences both stroke etiology and response to reperfusion therapies^1–4^. Thrombi have traditionally been classified as red blood cell (RBC)-rich (“red”) or fibrin/platelet-rich (“white”), reflecting low-versus high-flow thrombotic environments^2,5–8^, but this dichotomy has not been consistently confirmed in intracranial thrombi, where histopathology shows substantial overlap between etiological subtypes and marked intrathrombus heterogeneity^2,9–11^. Thrombus organization, particularly the distribution of platelet- and fibrin-rich regions between core and outer shell, may be more informative than the overall proportion of RBCs or fibrin. Platelet-rich outer layers enriched in fibrin and von Willebrand factor resist both thrombolysis and mechanical thrombectomy^12^, whereas RBC-rich regions are more susceptible to reperfusion. Consistently, greater platelet content is associated with higher thrombus stiffness, more severe stroke and worse revascularization outcomes, and platelet distribution has recently been proposed as a stronger classifier of thrombus phenotype than RBC or fibrin content alone^13^.

Immunothrombosis, the integration of innate immunity and coagulation^14,15^, is increasingly recognized as central to arterial occlusion. Neutrophil extracellular traps (NETs), extracellular chromatin associated with histones and granular proteins^16^, mediate thrombus formation by providing a scaffold for platelet adhesion, coagulation activation and fibrin stabilization^17–20^. Platelets in turn amplify NETosis through CD61-mediated and other platelet-neutrophil signaling pathways, generating a fibrin-NET network that confers thrombus stability and resistance to fibrinolysis^19,21,22^.

In human AIS thrombi, NETs are constitutive components across all etiological subtypes^23,24^, and their content correlates with endovascular procedure length and number of device passes^25–27^. The NET-rich outer shell contributes to resistance to both tPA-mediated thrombolysis and mechanical thrombectomy^12,28^, and DNase1, the principal NET-degrading enzyme, accelerates tPA-induced thrombolysis of retrieved cerebral thrombi^23,25,29–31^. Together, these findings support NETosis as a therapeutic target and highlight the need to define how circulating biomarkers relate to the local immunothrombotic composition of cerebral thrombi.

Most studies have analyzed plasma or intrathrombus markers in isolation, leaving two questions. First, we hypothesized that circulating NETosis biomarkers reflect systemic neutrophil activation and broad thrombus composition but only partially capture local immunothrombotic activity within the occlusive clot, a distinction that matters because these biomarkers are increasingly proposed for prognosis and for stratification in NET-targeted therapies^19^, yet their relationship with the local profile of retrieved thrombi remains insufficiently defined. Second, we hypothesized that intrathrombus NETosis markers, analyzed as a biological continuum rather than divided a priori into red, white, etiological or histological categories, would define clinically meaningful phenotypes associated with distinct dimensions of stroke outcome. We therefore performed an integrated analysis of circulating and intrathrombus NETosis markers in LVO-AIS patients treated with mechanical thrombectomy, evaluating associations with thrombus composition, collateral circulation, 3-month recovery and long-term survival.

## Methods

### Study design and population

This prospective observational cohort study (NETsTROKE, 2018–2023) enrolled consecutive adults with anterior-circulation LVO-AIS undergoing mechanical thrombectomy at a comprehensive stroke center. Eligible patients were ≥ 18 years with pre-stroke mRS ≤ 2 and LVO confirmed on CT or MR angiography. To limit confounding of immunothrombotic markers, we excluded patients on immunosuppressive therapy, with active infection in the preceding days, or with severe concomitant systemic disease (advanced renal or hepatic disease, active malignancy). Because biological sampling depended on research-team availability, not all treated patients could be sampled; the analyzed cohort comprises those in whom thrombus material was successfully retrieved and processed. Clinical, imaging and laboratory data were collected prospectively. The study was conceived as whole-cohort thrombus profiling: thrombi were not pre-classified into biological subgroups for the primary analyses. TOAST categories and histological architecture were retained for descriptive and sensitivity analyses, but the main biomarker phenotypes derive from cohort-wide median splits of quantitative immunohistochemistry. The study was approved by the institutional ethics committee (18/520 and 20/604) and conducted according to the Declaration of Helsinki and Good Clinical Practice; written informed consent was obtained from participants or legal representatives. STROBE recommendations for observational cohort studies were followed (Supplementary Item A)^32^.

### Clinical, vascular and imaging variables

NIHSS was assessed at admission and at 24 hours by trained neurologists. Collateral circulation was graded on baseline CT angiography using the Tan scale^33^ and dichotomized as good (Tan 2–3, filling > 50% of the middle cerebral artery territory) versus poor (Tan 0–1). Reperfusion was graded by the modified Thrombolysis in Cerebral Infarction (mTICI) scale. Because mTICI ≥ 2b was near-universal and carried negligible discriminating variance, complete reperfusion (mTICI 3) served as the procedural covariate in all multivariable models. Etiology was classified by TOAST criteria. Three-month outcome was assessed with the modified Rankin Scale; mRS ≤ 2 defined functional independence. All-cause mortality was recorded throughout follow-up (censoring date 30 September 2024); median follow-up in survivors was 1,250 days (IQR 930–1,839).

### Blood sampling and plasma biomarker quantification

Peripheral blood was collected at admission in EDTA tubes and processed to platelet-poor plasma by sequential centrifugation (1,000g, 10 min; then 2,000g, 15 min, 4°C); aliquots were stored at −80°C. Plasma biomarkers were quantified in duplicate by ELISA: cell-free DNA (cfDNA, Invitrogen P11496), neutrophil elastase (NE, Abcam ab119553), myeloperoxidase (MPO, R&D DMYE00B) and soluble CD40 ligand (Thermo Fisher BMS293INST). Admission complete blood counts were obtained on automated hematology analyzers. Plasma cfDNA was available in 90 of 160 patients (missingness analysis, Supplementary Table 1).

### Thrombus processing, histology and digital pathology

Retrieved thrombi were fixed in 4% paraformaldehyde, paraffin-embedded and cut into serial 3-µm sections. Hematoxylin–eosin staining was performed on representative sections and slides were digitized at high resolution. Two blinded observers classified thrombi by predominant subtype (RBC-rich, fibrin/platelet-rich, mixed) and by histological organization stage (fresh, lytic, organized), resolving discrepancies by consensus. Digital image analysis (ImageJ) within standardized regions of interest yielded percent area of erythrocytes, fibrin/platelets and nucleated cells, summing to 100% per section. The relatively high median fibrin/platelet area (85%) reflects digital quantification of thrombus tissue area excluding empty space and large vacuoles, consistent with single-center LVO thrombectomy series using comparable pipelines.

### Intrathrombus NETosis biomarkers

Adjacent serial sections underwent streptavidin–peroxidase immunohistochemistry with DAB chromogen after antigen retrieval (citrate buffer pH 6.0, approximately 95°C, 60 min). Primary antibodies were CitH3 (Abcam ab281584, 1:200), neutrophil elastase (Merck MABS461, 1:100) and CD61 (diagnostic-grade reagent in routine use by the institutional pathology service for immunohistochemical detection of platelet integrin β3, 1:50); negative controls omitted the primary antibody. Slides were analyzed in ImageJ by two blinded evaluators using color deconvolution (H DAB) and standardized thresholding; results are percent DAB-positive area within the thrombus region of interest. Each marker was dichotomized at the cohort median (CitH3 6.62%, NE 3.95%, CD61 23.09%) to define high and low phenotype groups for all subsequent analyses. Because biological-sample availability differed across analytical blocks, sample size varied by analysis: of the 160 patients, quantitative digital composition was available in 132, intrathrombus NETosis markers in 145 (CD61), 151 (NE) and 137 (CitH3), and admission plasma cfDNA in 90 (Supplementary Tables 1-3). Analyses were complete-case within each block, with sample sizes indicated in the corresponding tables and legends. Formal inter-observer agreement statistics were not pre-specified.

### Statistical analysis

Continuous variables are reported as mean ± SD or median [IQR], categorical as n (%). Groups were compared with Welch’s t-test or Mann–Whitney U and chi-square or Fisher’s exact test according to distribution and cell counts.

Spearman correlations across all 48 tested intrathrombus × plasma/CBC pairs were corrected using the Benjamini–Hochberg false discovery rate procedure (pFDR < 0.05). FDR correction was not applied to the within-phenotype bivariate comparisons of Tables 4–6, which are descriptive and contextualize the multivariable models; the latter constitute the primary inferential layer.

For each principal endpoint (good collateral circulation, mRS ≤ 2 at 3 months, follow-up mortality), logistic regression was reported with progressive adjustment: crude, then age, then NIHSS, then age plus NIHSS, then age plus NIHSS plus complete reperfusion. Because the proportional-hazards assumption was not met for complete reperfusion, mortality was analyzed by logistic regression with interval-specific estimates rather than a single Cox model. Stepwise reporting shows the trajectory of the marker effect across successive adjustment levels, reducing the risk that a single specification appears misleadingly favorable. Model performance was assessed by area under the ROC curve (AUC), and incremental discrimination of the marker beyond the clinical model was tested by DeLong’s method (ΔAUC) with two-sided p-value.

To determine whether the CD61–mRS association reflected a thrombus phenotype rather than an indirect consequence of more successful reperfusion, we performed a counterfactual mediation analysis using the parametric g-formula with mTICI 3 as candidate mediator, age and admission NIHSS as confounders, and bootstrap percentile confidence intervals (2,000 replicates) for the total, natural direct and natural indirect effects and the proportion mediated. It yields interpretable probability-scale contrasts and accommodates non-collapsibility of odds ratios^34,35^. A directed acyclic graph supporting the causal interpretation is provided in Figure 4F; formalized in Supplementary Item A.

Pre-specified sensitivity analyses included stratification by TOAST etiology (cardioembolic, atherothrombotic; Supplementary Table 4) and, for the neutrophil elastase–collateral association, direct adjustment for cardioembolic etiology (Supplementary Table 5). A missingness analysis for admission plasma cfDNA is reported in Supplementary Table 1.

Two-sided p < 0.05 was considered statistically significant. Analyses were performed using Python 3.12 (numpy, pandas, scipy, scikit-learn) with custom implementations of logistic regression, DeLong’s test and the counterfactual mediation g-formula.

### Data sharing statement

The data and analysis code that support the findings of this study are available from the corresponding author upon reasonable request, subject to institutional data-sharing agreements.

## Results

### Patient cohort and thrombus characteristics

Of 539 consecutive AIS patients screened, 160 with retrievable thrombus material, interpretable histology and complete clinical follow-up constituted the analytical cohort (Figure 1). Baseline clinical and procedural characteristics are summarized in Table 1. Strokes were severe (median admission NIHSS 18 [IQR 9–23]), reperfusion rates were high (mTICI ≥ 2b in 97.5%, mTICI 3 in 62%) and collateral circulation was good in 37.5% of patients. Over a median follow-up of 1,250 days in survivors, 52.3% achieved functional independence at 3 months and follow-up all-cause mortality was 38.8%.

**Figure 1.**
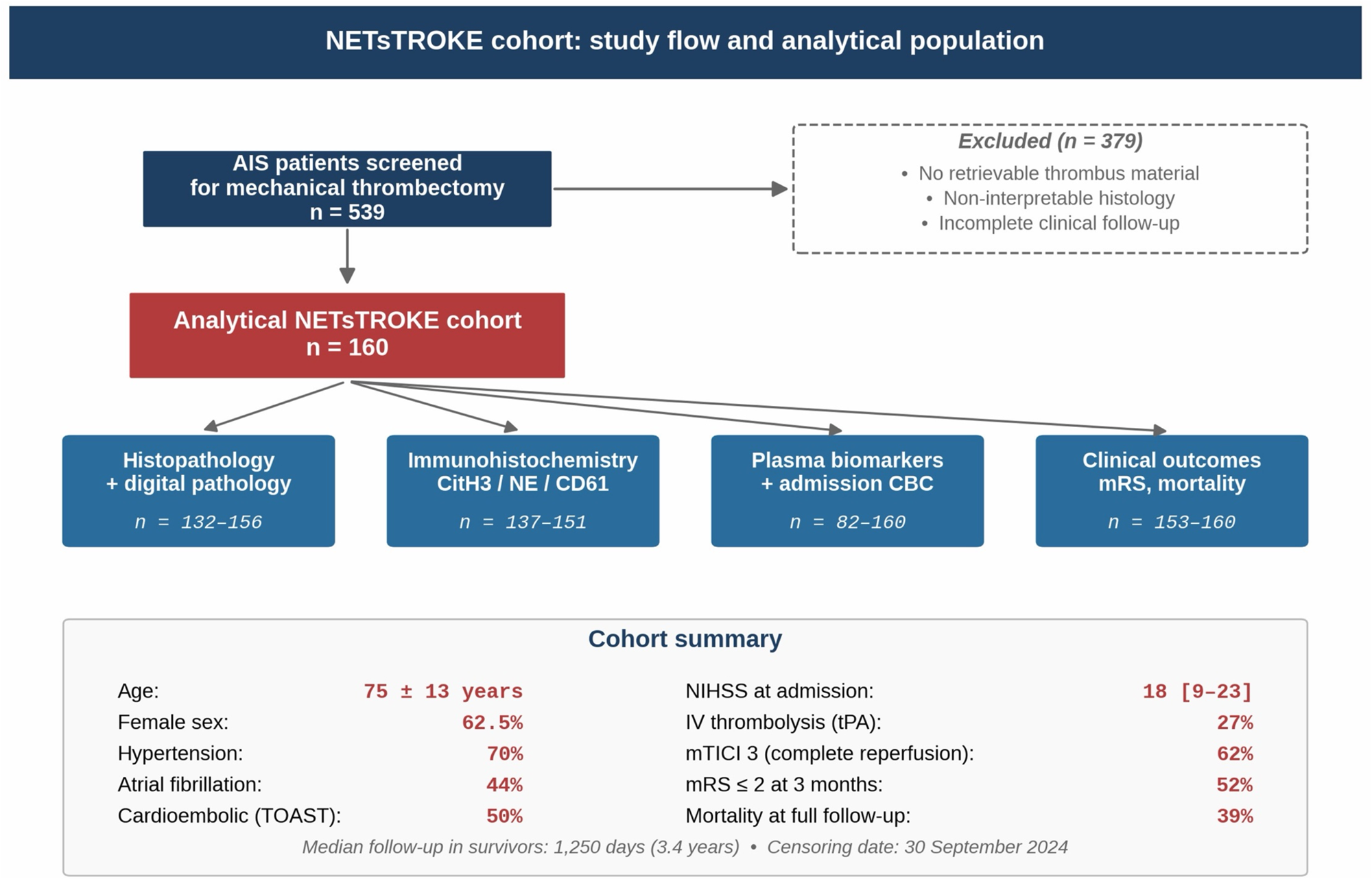
Cohort flow diagram and summary characteristics. Of 539 consecutive AIS patients screened, 160 with retrievable thrombus material and complete clinical follow-up constituted the analytical cohort. Demographic and procedural summary at the foot of the figure.

**Table 1.** Baseline characteristics of the NETsTROKE cohort (n = 160). TIA, transient ischaemic attack; NIHSS, National Institutes of Health Stroke Scale; ASPECTS, Alberta Stroke Program Early CT Score; mTICI, modified Thrombolysis in Cerebral Infarction; mRS, modified Rankin Scale; IQR, interquartile range. Censoring date:30 September 2024.

| Variable | Value | Missing,<br>n |
| --- | --- | --- |
| **Demographics** |  |  |
| Age, mean ± SD (years) | 75 ± 13 | 0 |
| Female sex, n (%) | 100 (62.5) | 0 |
| **Vascular risk factors, n (%)** |  |  |
| Hypertension | 112 (70.0) | 0 |
| Diabetes mellitus | 40 (25.0) | 0 |
| Dyslipidaemia | 96 (60.0) | 0 |
| Atrial fibrillation | 71 (44.4) | 0 |
| Previous stroke/TIA | 25 (15.6) | 0 |
| **Prior antithrombotic therapy, n (%)** |  |  |
| Antiplatelet | 36 (22.5) | 0 |
| Oral anticoagulation | 45 (28.1) | 0 |
| **Stroke etiology (TOAST), n (%)** |  |  |
| Cardioembolic | 80 (50.0) | 0 |
| Atherothrombotic | 34 (21.3) | 0 |
| Other/undetermined | 46 (28.7) | 0 |
| **Stroke severity and imaging** |  |  |
| NIHSS at admission, median [IQR] | 18 [9-23] | 0 |
| ASPECTS, median [IQR] | 9 [7-10] | 0 |
| **Treatment and procedural data** |  |  |
| Intravenous thrombolysis (tPA), n (%) | 43 (26.9) | 0 |
| Number of thrombectomy passes, median [IQR] | 2 [1-3] | 6 |
| mTICI ≥ 2b, n (%) | 156 (97.5) | 0 |
| mTICI 3 (complete reperfusion), n (%) | 99 (61.9) | 0 |
| **Outcomes** |  |  |
| Good collateral circulation (Tan 2-3), n (%) | 60 (37.5) | 0 |
| Functional independence at 3m (mRS ≤ 2), n (%) | 80 (52.3) | 7 |
| Follow-up mortality (all-cause), n (%) | 62 (38.8) | 0 |
| Median follow-up in survivors, days [IQR] | 1,250 [930-1,839] | 0 |

Retrieved thrombi were histopathologically heterogeneous (Figure 2C; Table 2): fibrin/platelet-rich thrombi predominated (43.9%), with mixed (29.0%) and RBC-rich (27.1%) patterns also represented. Digital pathology confirmed substantial inter-patient variability in RBC, fibrin/platelet and nucleated-cell content and heterogeneous expression of CitH3, NE and CD61 (Figure 2A–D). The three markers showed only modest co-distribution: CD61 correlated weakly with NE (ρ = +0.17, p = 0.049, n = 141), whereas neither correlated with CitH3, indicating that they capture partially distinct aspects of clot-associated immunothrombosis (Supplementary Table 2).

**Figure 2.**
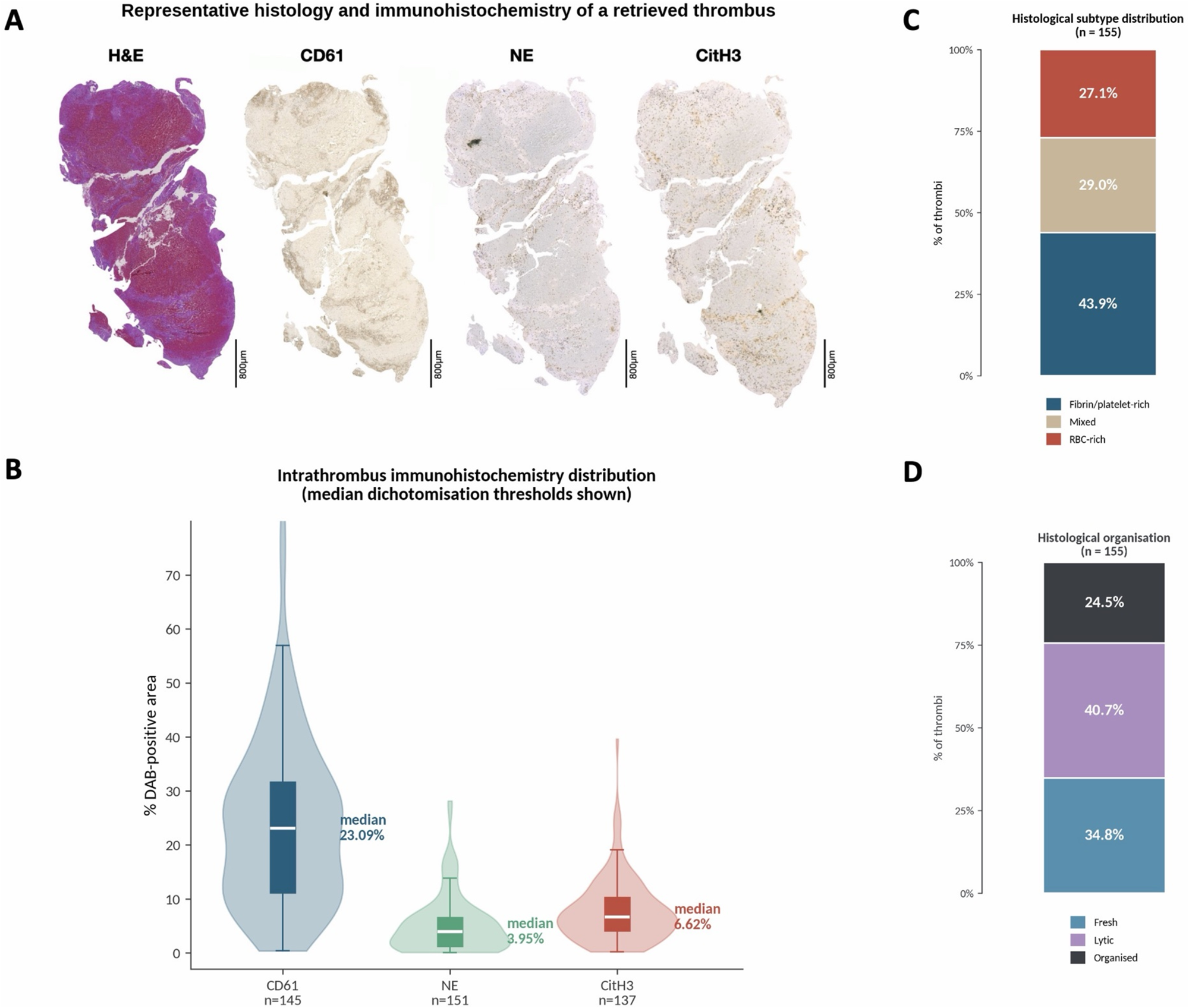
Histopathological and immunohistochemical characterization of retrieved thrombi. (A) Representative serial sections of a retrieved thrombus stained with H&E and DAB-IHC for CD61 (platelet integrin αIIbβ3), neutrophil elastase (NE) and citrullinated histone H3 (CitH3); scale bar 800 µm. (B) Distribution of intrathrombus % positive area for CD61, NE and CitH3 with median dichotomization thresholds indicated. (C) Distribution of histological subtypes (RBC-rich/mixed/fibrin-platelet-rich). (D) Distribution of histological organization (fresh/lytic/organized).

**Table 2.**
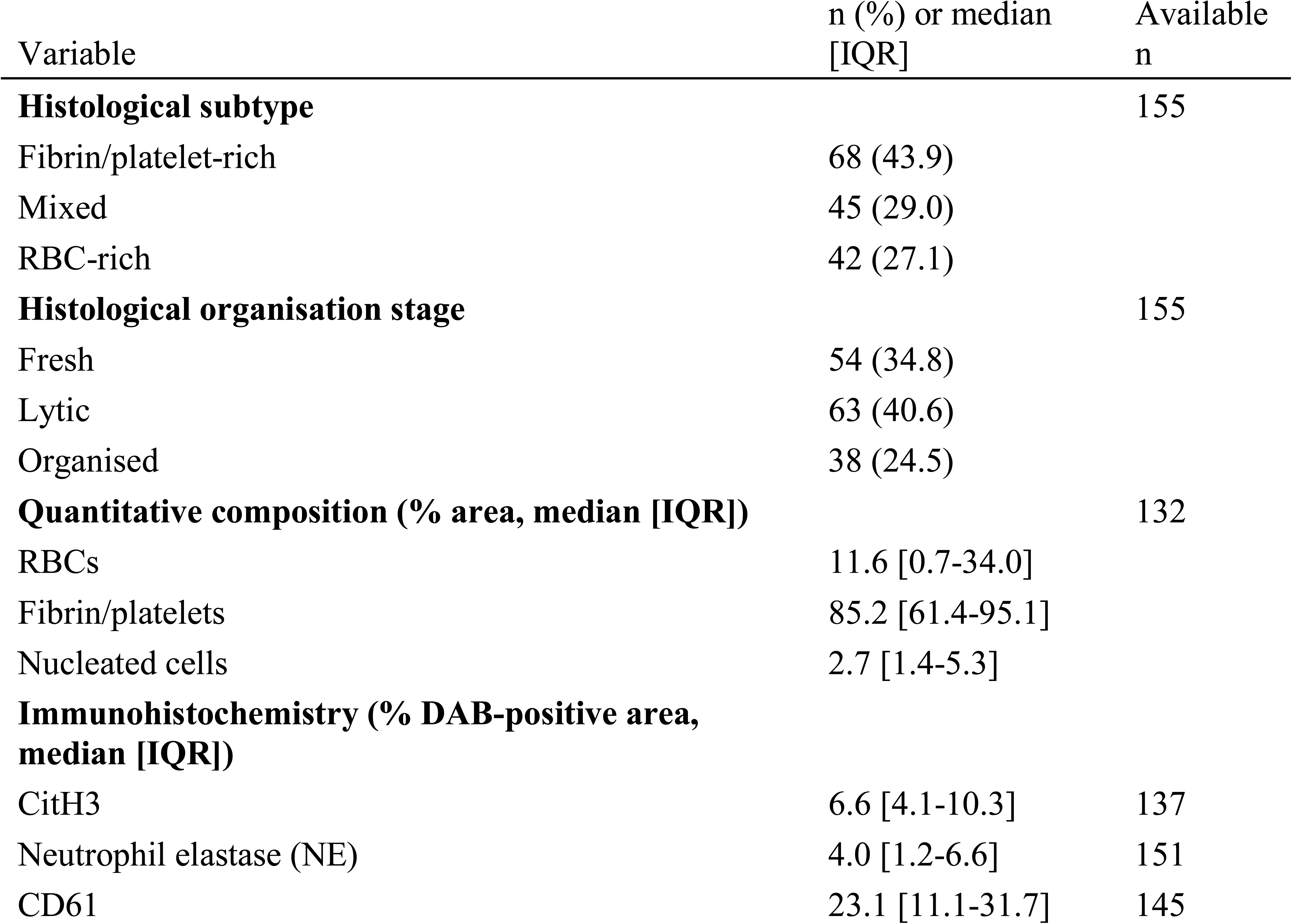
Histopathological characterization of retrieved thrombi. Median dichotomization cut-offs: CitH3 6.62%, NE 3.95%, CD61 23.09%.

### Circulating NETosis biomarkers reflect clot composition, but not the intrathrombus NETosis profile

We next asked whether admission plasma biomarkers could substitute for the local thrombus profile (Figure 3A-F; Table 3). Two patterns emerged. First, circulating NETosis biomarkers aligned with the macroscopic composition of the thrombus: RBC-rich thrombi correlated positively with plasma neutrophil elastase and MPO, whereas fibrin/platelet-rich thrombi correlated inversely with these markers and with circulating neutrophil and leukocyte counts, and all six architecture-related associations remained FDR-significant (Table 3). Within the plasma compartment, cfDNA correlated with neutrophil elastase (ρ = +0.31, pFDR = 0.008) and MPO (ρ = +0.29, pFDR = 0.011), which correlated strongly with each other (ρ = +0.69, pFDR < 0.001) and with sCD40L and circulating leukocyte and neutrophil counts, consistent with a coordinated systemic inflammatory and neutrophil-activation profile rather than a NETosis-specific signature.

**Figure 3.**
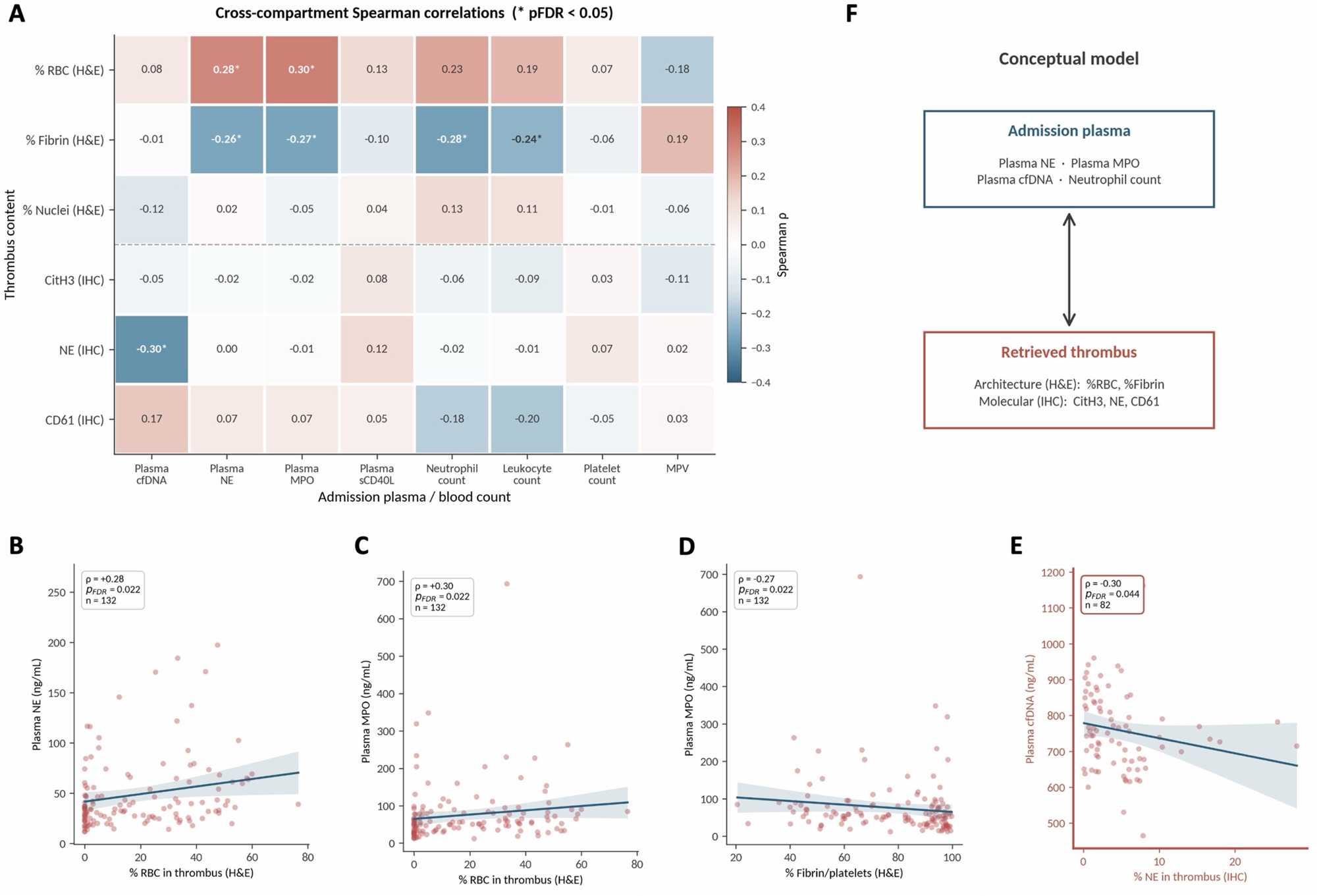
Cross-compartment plasma–thrombus correlation map. (A) Heatmap of Spearman correlations between thrombus variables (rows) and admission plasma/CBC variables (columns); asterisks denote FDR-significant pairs (Benjamini–Hochberg, pFDR < 0.05). (B–D) Scatter plots illustrating three of the six FDR-significant architectural-plasma pairs: %RBC vs plasma NE, %RBC vs plasma MPO, %fibrin/platelets vs plasma MPO. (E) The single FDR-significant cross-compartment IHC–plasma pair: inverse coupling between intrathrombus NE and plasma cfDNA. (F) Conceptual diagram summarizing architectural and molecular markers.

**Table 3.** FDR-significant cross-compartment correlations (Spearman). Benjamini–Hochberg correction across 48 tested intrathrombus × plasma/CBC pairs. None of the intrathrombus IHC markers (CitH3, NE, CD61) showed a significant positive correlation with their respective plasma counterparts after FDR correction.

| Thrombus variable | Plasma/CBC variable | $\rho$ | pFDR |
| --- | --- | --- | --- |
| **Architecture aligned with plasma surrogates and circulating leukocytes** |  |  |  |
| % RBC (H&E) | Plasma neutrophil elastase | +0.28 | **0.022** |
| % RBC (H&E) | Plasma MPO | +0.30 | **0.022** |
| % Fibrin/platelets (H&E) | Plasma neutrophil elastase | -0.26 | **0.022** |
| % Fibrin/platelets (H&E) | Plasma MPO | -0.27 | **0.022** |
| % Fibrin/platelets (H&E) | Neutrophil count ($\times 10^9/L$) | -0.28 | **0.022** |
| % Fibrin/platelets (H&E) | Leukocyte count ($\times 10^9/L$) | -0.24 | **0.044** |
| **Coordinated systemic NETosis signature centred on cfDNA** |  |  |  |
| Plasma cfDNA | Plasma neutrophil elastase | +0.31 | **0.008** |
| Plasma cfDNA | Plasma MPO | +0.29 | **0.011** |
| Plasma neutrophil elastase | Plasma MPO | +0.69 | **<0.001** |
| Plasma neutrophil elastase | Plasma sCD40L | +0.29 | **0.001** |
| Plasma MPO | Plasma sCD40L | +0.23 | **0.008** |
| Plasma neutrophil elastase | Neutrophil count ($\times 10^9/L$) | +0.24 | **0.005** |
| Plasma neutrophil elastase | Leukocyte count ($\times 10^9/L$) | +0.27 | **0.002** |
| Plasma MPO | Neutrophil count ($\times 10^9/L$) | +0.25 | **0.005** |
| Plasma MPO | Leukocyte count ($\times 10^9/L$) | +0.28 | **0.001** |
| **Cross-compartment IHC-plasma dissociation (only FDR-significant pair)** |  |  |  |
| Intrathrombus NE (% area) | Plasma cfDNA | **-0.30** | **0.044** |

Second, this systemic coherence did not extend to the thrombus. The only FDR-significant cross-compartment association was an inverse correlation between intrathrombus NE and plasma cfDNA (ρ = −0.30, pFDR = 0.044), and no intrathrombus marker correlated positively with its circulating counterpart (full 48-pair matrix, Supplementary Table 3). The plasma signal therefore indexed broad clot architecture, whereas the outcome-associated intrathrombus phenotypes were largely independent of circulating NETosis readouts.

### High intrathrombus CD61 identifies a platelet/fibrin-enriched phenotype associated with functional independence

Patients dichotomized into low (n = 73) and high (n = 72) intrathrombus CD61 differed in thrombus architecture, stroke severity and clinical outcome (Figure 4A-F; Table 4). High-CD61 thrombi were more often fibrin/platelet-rich and less often RBC-rich, supporting CD61 as a marker of a platelet/fibrin-enriched phenotype rather than a surrogate of total clot cellularity. Clinically, high CD61 was associated with lower C-reactive protein (p = 0.033), better preserved renal function (p = 0.028), lower admission NIHSS (15.1 ± 7.7 vs 17.8 ± 6.4; p = 0.027), fewer tandem occlusions (14% vs 29%; p = 0.047), better collateral circulation (44% vs 27%; p = 0.038), higher 3-month functional independence (mRS ≤ 2: 62.5% vs 39.7%; p = 0.008) and lower follow-up mortality.

**Figure 4.**
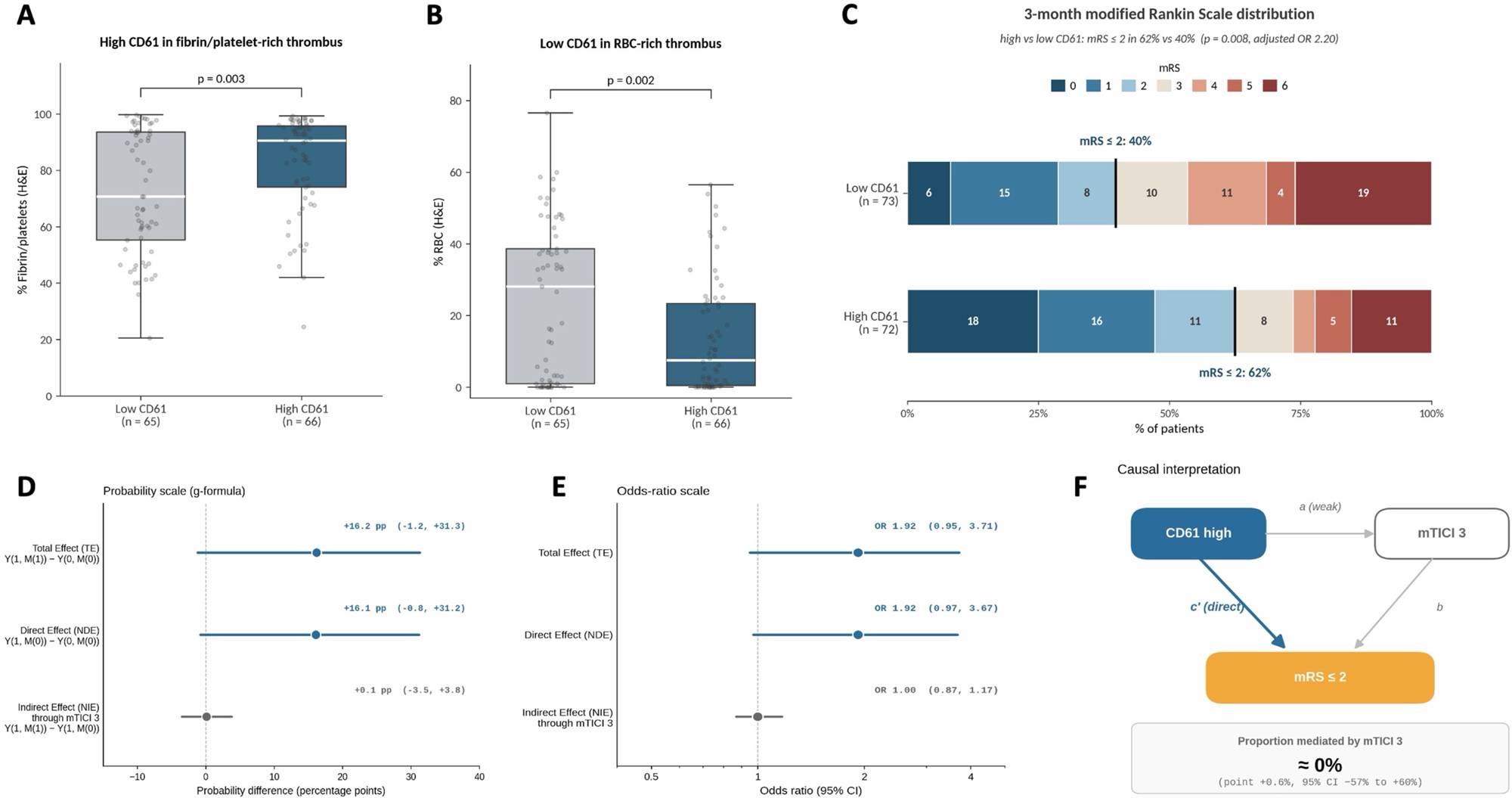
CD61: a fibrin-organized phenotype with favorable functional outcome independent of complete reperfusion. (A, B) Box plots of intrathrombus architecture (%fibrin/platelets and %RBC) by CD61 high vs low. (C) Stacked bar of 3-month mRS distribution by CD61 high vs low (mRS ≤ 2:62.5 vs 39.7%, p = 0.008). (D, E) Counterfactual mediation analysis (parametric g-formula): forest plot of the total effect (TE), natural direct effect (NDE) and natural indirect effect (NIE) of high CD61 on the probability of mRS ≤ 2, with OR-scale equivalents. The natural indirect effect through complete reperfusion (mTICI 3) is essentially zero (NIE +0.001 probability units; OR-NIE 1.00, 95% CI 0.87-1.17), supporting a direct phenotypic association rather than a procedural proxy. (F) Causal interpretation.

**Table 4.**
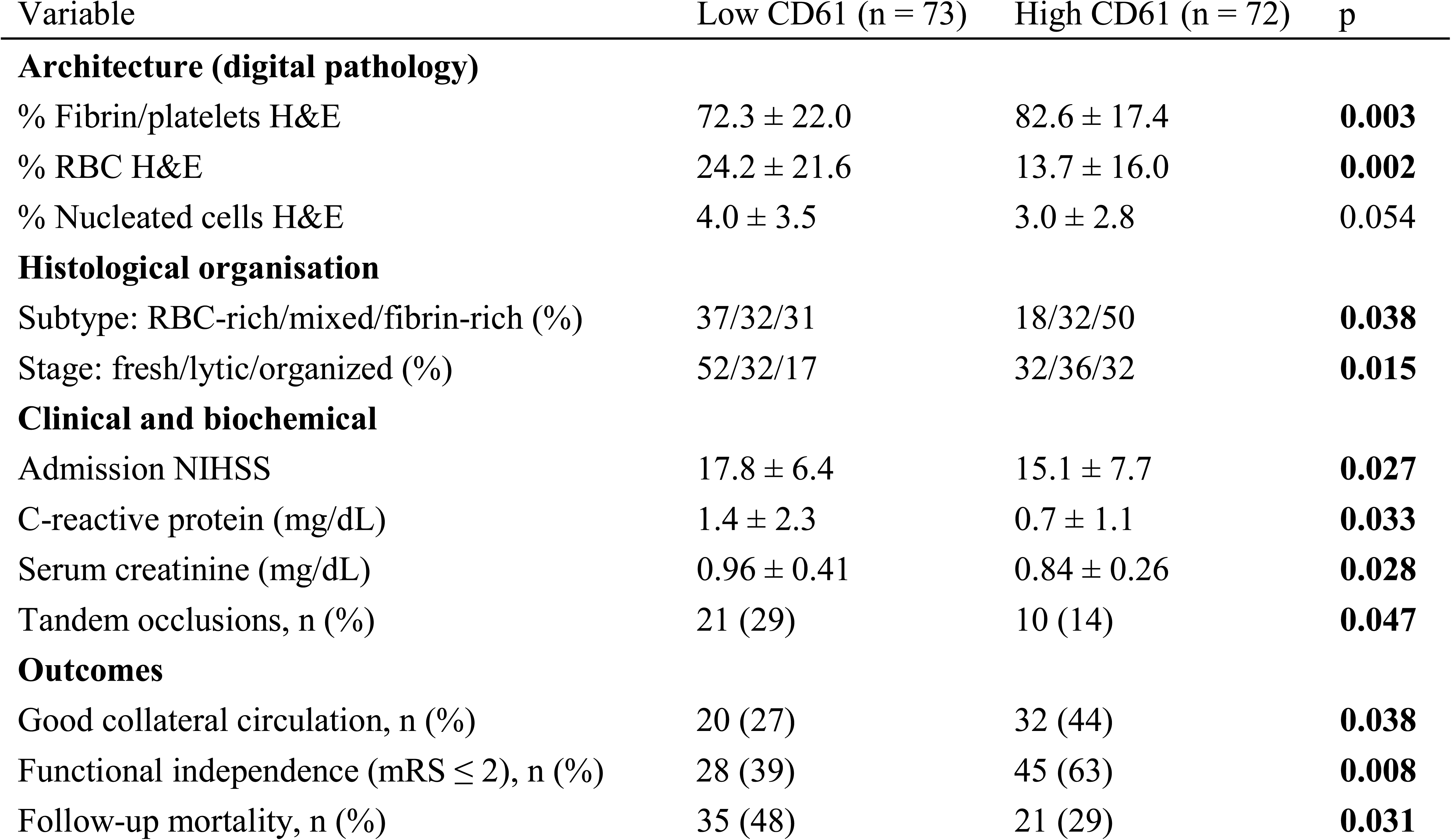
CD61 phenotype: multidimensional characterization. Continuous variables: mean ± SD, Welch’s t-test. Categorical: chi-square or Fisher’s exact test. Multivariable trajectories in Table 7.

| Variable | Low CD61 (n = 73) | High CD61 (n = 72) | p |
| --- | --- | --- | --- |
| **Architecture (digital pathology)** |  |  |  |
| % Fibrin/platelets H&E | 72.3 ± 22.0 | 82.6 ± 17.4 | **0.003** |
| % RBC H&E | 24.2 ± 21.6 | 13.7 ± 16.0 | **0.002** |
| % Nucleated cells H&E | 4.0 ± 3.5 | 3.0 ± 2.8 | 0.054 |
| **Histological organisation** |  |  |  |
| Subtype: RBC-rich/mixed/fibrin-rich (%) | 37/32/31 | 18/32/50 | **0.038** |
| Stage: fresh/lytic/organized (%) | 52/32/17 | 32/36/32 | **0.015** |
| **Clinical and biochemical** |  |  |  |
| Admission NIHSS | 17.8 ± 6.4 | 15.1 ± 7.7 | **0.027** |
| C-reactive protein (mg/dL) | 1.4 ± 2.3 | 0.7 ± 1.1 | **0.033** |
| Serum creatinine (mg/dL) | 0.96 ± 0.41 | 0.84 ± 0.26 | **0.028** |
| Tandem occlusions, n (%) | 21 (29) | 10 (14) | **0.047** |
| **Outcomes** |  |  |  |
| Good collateral circulation, n (%) | 20 (27) | 32 (44) | **0.038** |
| Functional independence (mRS ≤ 2), n (%) | 28 (39) | 45 (63) | **0.008** |
| Follow-up mortality, n (%) | 35 (48) | 21 (29) | **0.031** |

In progressively adjusted models, high CD61 remained independently associated with 3-month functional independence after adjustment for age, admission NIHSS and complete reperfusion, with a consistent effect size across all adjustment levels (Table 7, panel A). We therefore tested by mediation analysis whether this association was explained by higher rates of complete reperfusion. The indirect effect through mTICI 3 was minimal and the direct effect of CD61 remained largely unchanged (Figure 4D), indicating that the association is not primarily driven by procedural success but reflects a broader biological phenotype. Incremental discrimination beyond the clinical model was modest (ΔAUC = +0.013, DeLong p = 0.47)(TOAST-stratified analysis, Supplementary Table 4).

### High intrathrombus NE is associated with favorable collateral circulation

Patients dichotomized into low (n = 76) and high (n = 75) intrathrombus NE showed a more focused biological and clinical profile (Figure 5A-C; Table 5). Plasma cfDNA was lower in high-NE patients, consistent with the inverse correlation above, and high NE was associated with higher intrathrombus CD61 (p = 0.027) but not CitH3. High NE was associated with good collateral circulation (45.3% vs 26.3%; p = 0.018) but not with 3-month functional independence or mortality. In progressively adjusted models this association persisted after adjustment for age, admission NIHSS and complete reperfusion and was maintained after additional adjustment for cardioembolic etiology (adjusted OR 2.90, 95% CI 1.39–6.08; p = 0.005; Table 7, panel B, Supplementary Table 5). Incremental discrimination was modest (ΔAUC = +0.040, DeLong p = 0.25).

**Figure 5.**
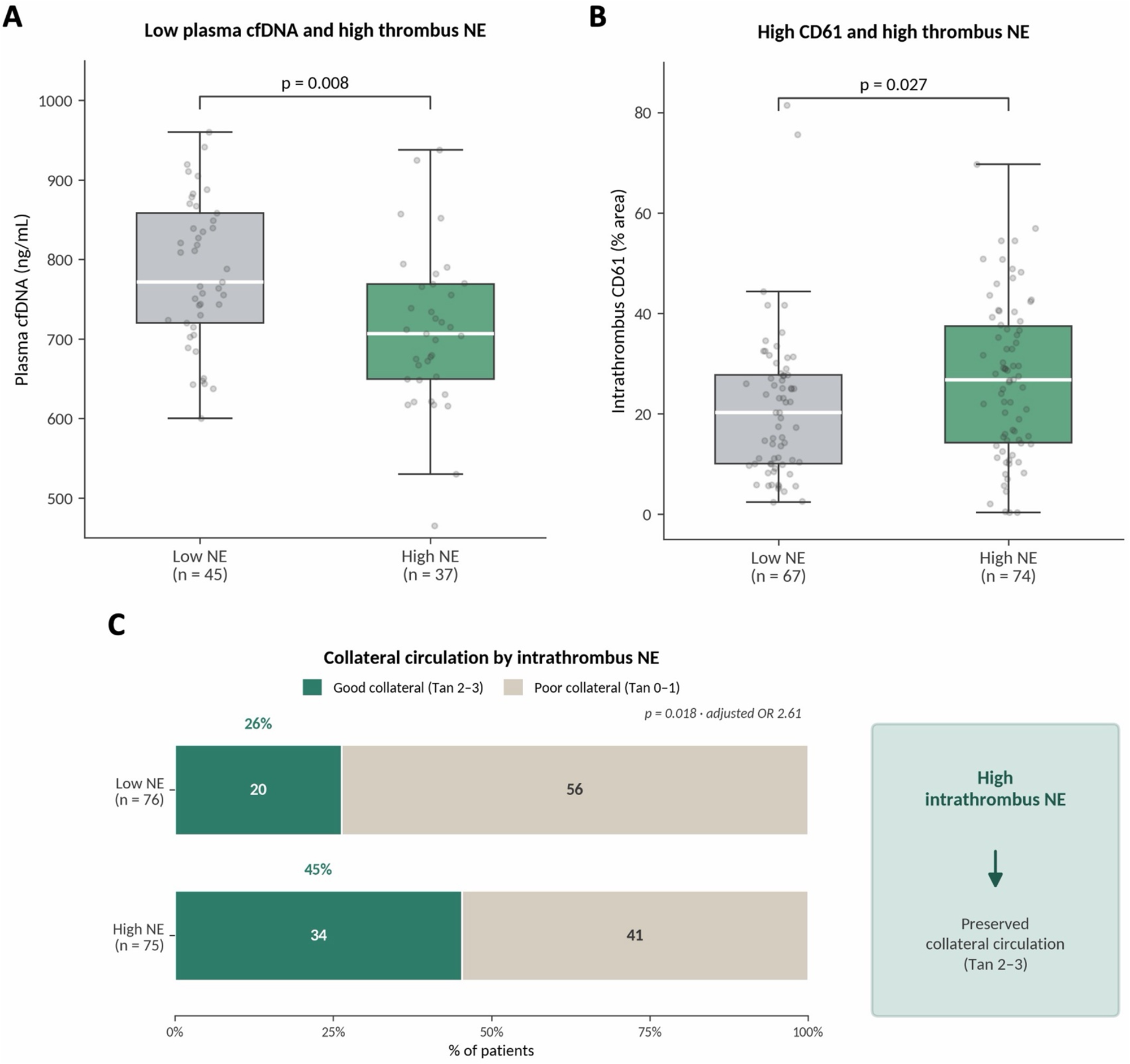
NE: independent correlate of preserved collateral circulation. (A) Plasma cfDNA by NE high vs low (722 vs 791 ng/mL, p = 0.008). (B) Intrathrombus CD61 by NE high vs low (26.7 vs 21.0%, p = 0.027). (C) Stacked bar of good collateral circulation by NE high vs low (45.3 vs 26.3%, p = 0.018).

**Table 5.** NE phenotype: multidimensional characterization. Continuous variables: mean ± SD, Welch’s t-test. Categorical: chi-square or Fisher’s exact test. Multivariable trajectories in Table 7.

| Variable | Low NE (n = 76) | High NE (n = 75) | p |
| --- | --- | --- | --- |
| **Plasma (cross-compartment dissociation)** |  |  |  |
| Plasma cfDNA, ng/mL (n = 82 paired) | 791 ± 109 | 722 ± 120 | **0.008** |
| Plasma neutrophil elastase, ng/mL | 45.9 ± 26.0 | 51.3 ± 39.4 | 0.32 |
| Plasma MPO, ng/mL | 21.0 ± 11.5 | 23.2 ± 14.7 | 0.31 |
| **Intrathrombus convergence** |  |  |  |
| Intrathrombus CD61 (% area) | 21.0 ± 14.6 | 26.7 ± 15.4 | **0.027** |
| Intrathrombus CitH3 (% area) | 5.4 ± 5.9 | 5.6 ± 5.9 | 0.86 |
| **Architecture (digital pathology)** |  |  |  |
| % Fibrin/platelets H&E | 74.4 ± 21.0 | 80.1 ± 19.7 | 0.110 |
| % RBC H&E | 22.2 ± 20.2 | 16.2 ± 18.9 | 0.083 |
| **Outcome** |  |  |  |
| Good collateral circulation, n (%) | 20 (27) | 34 (45) | **0.018** |
| Functional independence (mRS ≤ 2), n (%) | 38 (52.8) | 38 (52.8) | 1.00 |
| Follow-up mortality, n (%) | 34 (44.7) | 27 (36.0) | 0.32 |

**Table 6.**
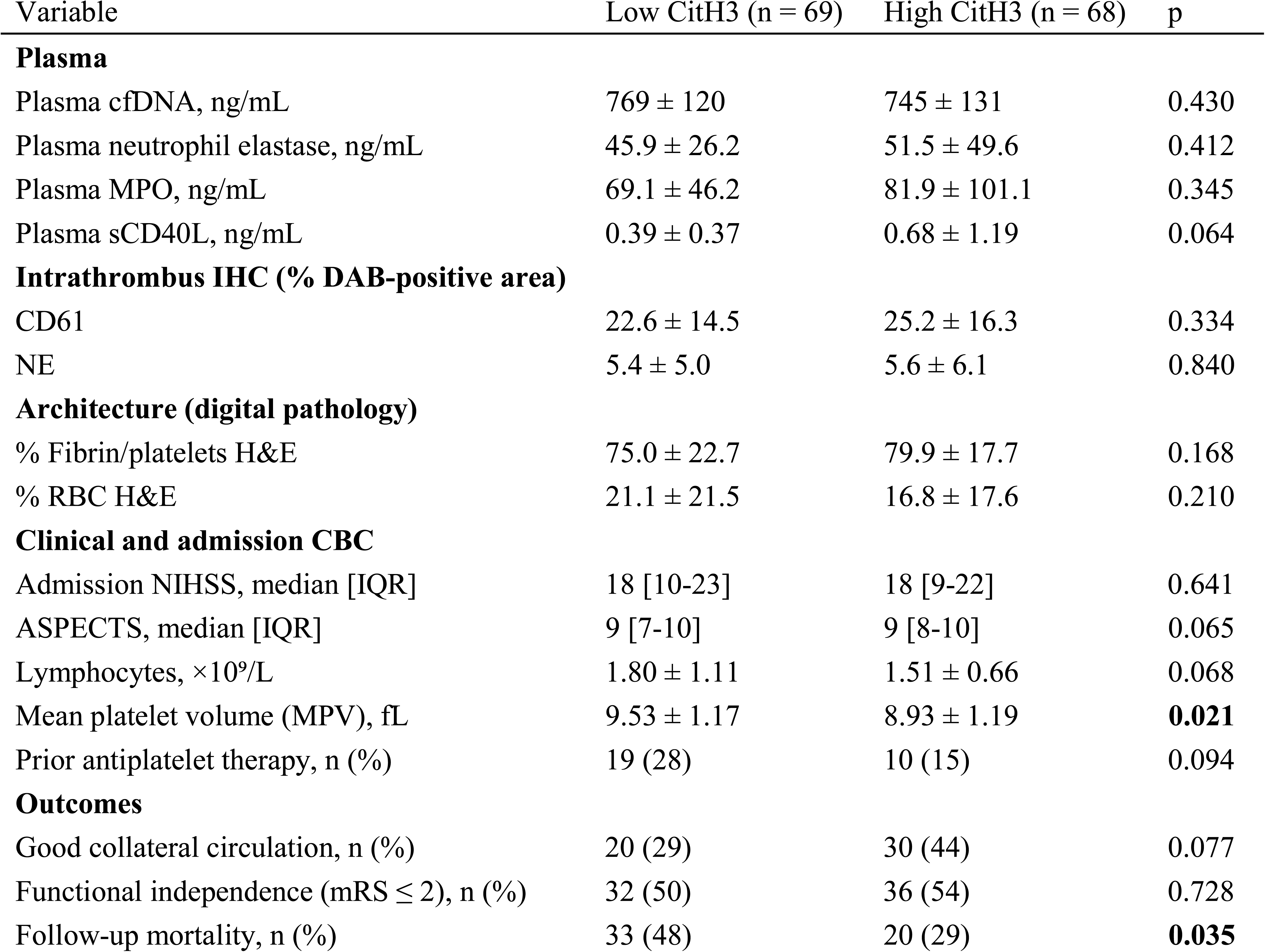
CitH3 phenotype: multidimensional characterization. Continuous variables: mean ± SD, Welch’s t-test. Categorical: chi-square or Fisher’s exact test. Multivariable trajectories in Table 7.

| Variable | Low CitH3 (n = 69) | High CitH3 (n = 68) | p |
| --- | --- | --- | --- |
| **Plasma** |  |  |  |
| Plasma cfDNA, ng/mL | 769 ± 120 | 745 ± 131 | 0.430 |
| Plasma neutrophil elastase, ng/mL | 45.9 ± 26.2 | 51.5 ± 49.6 | 0.412 |
| Plasma MPO, ng/mL | 69.1 ± 46.2 | 81.9 ± 101.1 | 0.345 |
| Plasma sCD40L, ng/mL | 0.39 ± 0.37 | 0.68 ± 1.19 | 0.064 |
| **Intrathrombus IHC (% DAB-positive area)** |  |  |  |
| CD61 | 22.6 ± 14.5 | 25.2 ± 16.3 | 0.334 |
| NE | 5.4 ± 5.0 | 5.6 ± 6.1 | 0.840 |
| **Architecture (digital pathology)** |  |  |  |
| % Fibrin/platelets H&E | 75.0 ± 22.7 | 79.9 ± 17.7 | 0.168 |
| % RBC H&E | 21.1 ± 21.5 | 16.8 ± 17.6 | 0.210 |
| **Clinical and admission CBC** |  |  |  |
| Admission NIHSS, median [IQR] | 18 [10-23] | 18 [9-22] | 0.641 |
| ASPECTS, median [IQR] | 9 [7-10] | 9 [8-10] | 0.065 |
| Lymphocytes, ×10<sup>9</sup>/L | 1.80 ± 1.11 | 1.51 ± 0.66 | 0.068 |
| Mean platelet volume (MPV), fL | 9.53 ± 1.17 | 8.93 ± 1.19 | **0.021** |
| Prior antiplatelet therapy, n (%) | 19 (28) | 10 (15) | 0.094 |
| **Outcomes** |  |  |  |
| Good collateral circulation, n (%) | 20 (29) | 30 (44) | 0.077 |
| Functional independence (mRS ≤ 2), n (%) | 32 (50) | 36 (54) | 0.728 |
| Follow-up mortality, n (%) | 33 (48) | 20 (29) | **0.035** |

**Table 7.** Multivariable logistic regression: progressive adjustment for the three principal endpoints. OR, odds ratio; CI, confidence interval; AUC, area under the receiver-operating-characteristic curve; ΔAUC, incremental discrimination of the marker beyond the corresponding base clinical model, tested by DeLong’s method. Each table reports the same patients across all rows (complete-case in the listed n).

**A. Intrathrombus CD61 → 3-month functional independence (mRS ≤ 2). n = 145, events = 74.**
| Adjustment level | OR (95% CI) | p | AUC |
| --- | --- | --- | --- |
| Crude | 2.53 (1.30-4.94) | 0.007 | 0.614 |
| + age | 2.54 (1.29-5.02) | 0.007 | 0.661 |
| + NIHSS | 2.13 (1.05-4.34) | 0.037 | 0.728 |
| + age + NIHSS | 2.13 (1.03-4.38) | 0.040 | 0.744 |
| **+ age + NIHSS + mTICI 3** | **2.20 (1.05-4.60)** | **0.036** | **0.762** |

**B. Intrathrombus NE → good collateral circulation. n = 151, events = 54.**
| Adjustment level | OR (95% CI) | p | AUC |
| --- | --- | --- | --- |
| Crude | 2.32 (1.17-4.60) | 0.016 | 0.603 |
| + age | 2.32 (1.17-4.61) | 0.016 | 0.608 |
| + NIHSS | 2.61 (1.28-5.33) | 0.009 | 0.688 |
| + age + NIHSS | 2.62 (1.28-5.36) | 0.008 | 0.688 |
| **+ age + NIHSS + mTICI 3** | **2.61 (1.28-5.35)** | **0.009** | **0.687** |
| Sensitivity: + cardioembolic etiology (n = 151, events = 54; full adjustment) | **2.90 (1.39-6.08)** | **0.005** | **0.694** |

**C. Intrathrombus CitH3 → follow-up all-cause mortality. n = 137, events = 53.**
| Adjustment level | OR (95% CI) | p | AUC |
| --- | --- | --- | --- |
| Crude | 0.45 (0.22-0.92) | 0.028 | 0.597 |
| + age | 0.38 (0.18-0.83) | 0.015 | 0.738 |
| + NIHSS | 0.44 (0.21-0.92) | 0.029 | 0.703 |
| + age + NIHSS | 0.36 (0.16-0.80) | 0.012 | 0.781 |
| **+ age + NIHSS + mTICI 3** | **0.33 (0.14-0.74)** | **0.008** | **0.785** |

ΔAUC: +0.023 (DeLong p = 0.26).

### High intrathrombus CitH3 is associated with lower mortality

Dichotomized at the median (low, n = 69; high, n = 68), high intrathrombus CitH3 was associated with lower mean platelet volume at admission (p = 0.021) and lower long-term all-cause mortality (29% vs 48%; p = 0.035) (Figure 6A-D; Table 6). The association with reduced mortality remained significant after adjustment for age, admission NIHSS and complete reperfusion (Table 7, panel C), with modest incremental discrimination (ΔAUC = +0.023, DeLong p = 0.26). A pre-specified analysis across non-overlapping intervals showed the mortality difference to be most pronounced 3–5 years after stroke (Figure 6B; Supplementary Table 6); because events per interval were limited, these analyses are exploratory.

**Figure 6.**
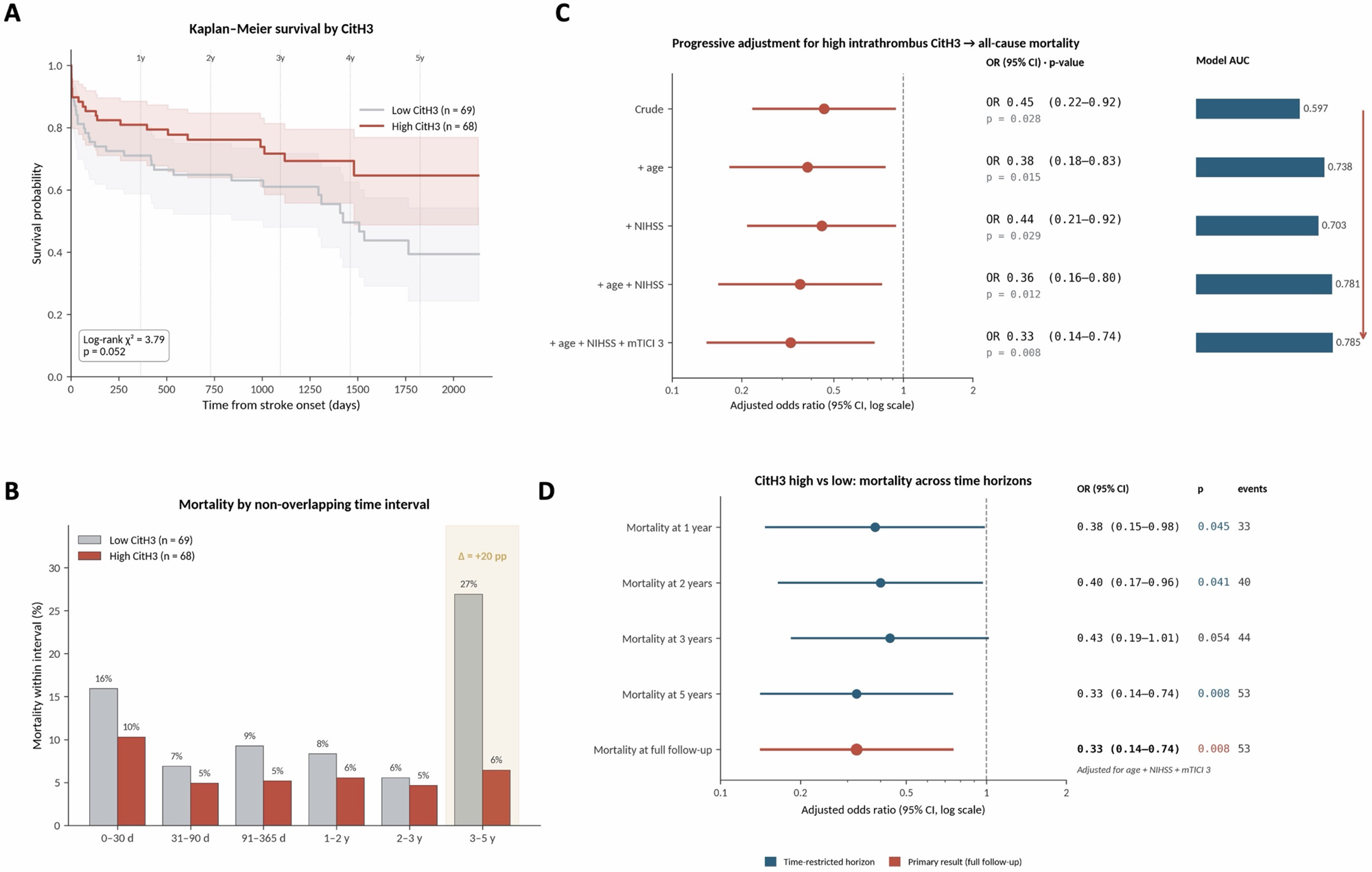
CitH3: long-term mortality association. (A) Kaplan–Meier survival curves for CitH3 high vs low across full follow-up. (B) Mortality by non-overlapping time intervals; the largest absolute difference occurs in the 3-5-year window (27 vs 6%, exploratory). (C) Forest plot of progressively adjusted logistic regression models. (D) Forest plot of CitH3 high vs low across multiple time horizons (1, 2, 3, 5 years and full follow-up).

### Intrathrombus NETosis markers show complementary, outcome-specific associations

To determine whether the three markers represented a single co-linear signal or independent prognostic information, we modelled CitH3, NE and CD61 simultaneously, adjusted for age and admission NIHSS (Table 8; a model additionally adjusted for complete reperfusion is provided in Supplementary Table 7). Despite modest inter-marker correlations, each showed preferential associations with distinct clinical outcomes: CD61 with 3-month functional independence, NE with good collateral circulation and CitH3 with reduced follow-up mortality. CD61, NE and CitH3 are therefore not interchangeable proxies of NET burden, but partially independent thrombus signatures with outcome-specific associations. Marker-endpoint associations were materially unchanged after additional adjustment for cardioembolic etiology, and TOAST etiology was not itself independently associated with any endpoint: comparing cardioembolic with non-cardioembolic strokes, 3-month functional independence was 48% versus 57% (p = 0.33) and good collateral circulation 34% versus 41% (p = 0.41); although crude follow-up mortality was numerically higher in cardioembolic strokes (46% versus 31%, p = 0.07), this difference did not persist after adjustment for age and admission NIHSS (p = 0.29)(Supplementary Tables 4 and 5).

**Table 8.** Convergent multivariable model: each outcome is preferentially predicted by a different intrathrombus marker. Each model included all three intrathrombus markers simultaneously and was adjusted for age and admission NIHSS (mTICI 3 omitted from this convergent table due to near-quasi-complete separation in two of the three outcomes; full model with mTICI 3 in Supplementary Table S5). Statistically significant associations (p < 0.05) are shown in bold.

| Outcome (n = 132) | Predictor | OR (95% CI) | p | AUC |
| --- | --- | --- | --- | --- |
| **Good collateral circulation** (events = 46) | NE high | **2.54 (1.13-5.71)** | 0.024 | 0.732 |
|  | CitH3 high | **2.28 (1.04-5.01)** | 0.040 |  |
|  | CD61 high | 1.88 (0.85-4.14) | 0.118 |  |
| **Functional independence (mRS ≤ 2)** (events = 68) | CD61 high | 1.98 (0.91-4.30) | 0.085 | 0.758 |
|  | NE high | 1.03 (0.47-2.26) | 0.939 |  |
|  | CitH3 high | 1.18 (0.55-2.55) | 0.671 |  |
| **Follow-up mortality** (events = 52) | CitH3 high | **0.32 (0.14-0.75)** | 0.009 | 0.797 |
|  | CD61 high | 0.51 (0.22-1.18) | 0.115 |  |
|  | NE high | 0.67 (0.29-1.55) | 0.349 |  |

## Discussion

In this prospective cohort of 160 patients with LVO-AIS treated with mechanical thrombectomy, intrathrombus NETosis-related markers defined prognostically distinct phenotypes, plasma biomarkers primarily reflected macroscopic RBC-fibrin composition rather than the local profile, and analysing all thrombi as a unified cohort allowed quantitative median thresholds to identify phenotypes cutting across conventional red/white and etiological categories. None of the three phenotypes was explained by stroke etiology: associations were independent of cardioembolic versus non-cardioembolic classification, and TOAST subtype showed no independent association with functional outcome, collateral status or survival, indicating that quantitative immunothrombotic composition conveys prognostic information not reducible to established subtypes. Whereas prior work emphasized etiology, subtypes, aggregate NET burden or outcome-optimized cutoffs, we treat retrieved thrombi as a single quantitative biomarker space, making thresholds transparent and reproducible although exploratory until externally validated.

### Plasma and intrathrombus NETosis capture complementary, non-interchangeable dimensions of immunothrombosis

Circulating NE, MPO and leukocyte counts tracked the RBC-fibrin composition of retrieved thrombi, but this did not translate into correspondence with local NETosis: after correction for multiple testing the only significant cross-compartment association was an inverse correlation between plasma cfDNA and intrathrombus NE, and no intrathrombus marker correlated positively with its circulating counterpart. Circulating biomarkers are therefore informative for broad composition but not interchangeable with thrombus-level profiling, consistent with a previous prospective thrombectomy cohort^36,37^. The inference is tempered by the limited NETosis specificity of cfDNA, which reflects total cell-free DNA from multiple sources; the single cross-compartment association therefore rests on the least NET-specific analyte and warrants confirmation with MPO-DNA or CitH3-DNA complexes, and further relationships may emerge in larger studies.

Several mechanisms may explain this discordance. The fibrin-platelet outer shell may retain NET-derived material within the clot^12,38^, limiting its release; conversely, elevated circulating biomarkers may reflect diffuse neutrophil activation, NET spillover or impaired degradation rather than greater local accumulation, consistent with studies linking higher cfDNA and MPO-DNA complexes and reduced DNase activity to greater severity and worse outcome^19,39,40^. Comparable discrepancies have been reported in coronary and cerebral thrombi^36,41–43^. Spatially resolved approaches such as multiplex immunofluorescence and spatial transcriptomics will be needed to define how NETosis is distributed within the thrombus.

### CD61-rich thrombi and functional recovery

The CD61-enriched phenotype combined better collaterals, lower baseline severity, higher functional independence and lower mortality, and remained stable across sequential adjustment; mediation showed only a minimal indirect effect through mTICI 3. It may appear paradoxical, since platelet-rich thrombi have been linked to reperfusion resistance and poorer revascularization^2,44,45^. However, CD61 immunohistochemistry reflects platelet antigen presence, including quiescent platelets and platelet-containing fibrin networks, and does not measure activation or ongoing aggregation; a stronger signal may therefore mark a structurally organized platelet/fibrin phenotype rather than an actively prothrombotic state. High CD61 was also associated with better preserved renal function (creatinine 0.84 versus 0.96 mg/dL; p = 0.028), raising the possibility that uremia-related platelet dysfunction influences CD61 immunoreactivity, but the association with functional independence persisted after adjustment for creatinine. These data do not prove CD61 is protective; they indicate it enriches for a favourable thrombus-patient phenotype distinct from circulating biomarkers and conventional platelet-rich classifications. Markers of platelet activation such as P-selectin or activated GPIIb/IIIa, with spatial profiling, will be needed to establish whether this reflects platelet quiescence, structural organization, vascular reserve or a combination.

### NE-rich thrombi and collateral circulation

High intrathrombus NE was independently linked to favourable collateral circulation across progressively adjusted models and remained robust after direct adjustment for cardioembolic etiology (adjusted OR 2.90, 95% CI 1.39-6.08; p = 0.005), indicating it does not merely reflect the poorer baseline collateral status typical of cardioembolic strokes. It did not translate into better functional outcome or lower mortality, suggesting that thrombus NE relates more closely to vascular reserve than to the overall post-stroke trajectory. NE may reflect the activation state of neutrophils at occlusion rather than acting only as a tissue-damaging mediator: neutrophils with preserved degranulation capacity may retain vascular regulatory functions in collateral adaptation and endothelial homeostasis^46^. Consistent with the limited cross-compartment concordance, the inverse relationship between thrombus NE and plasma cfDNA indicates that local enrichment does not imply greater systemic NET release. We recently showed that DNase1 functional capacity, influenced by the rs1053874 polymorphism, modulates circulating NET burden and is associated with early neurological recovery and long-term survival^40^. Experimental and clinical data support this reading: in murine circadian models, NETosis during the inactive phase promoted microvascular obstruction and impaired collateral perfusion, whereas inhibiting NET formation or deleting PAD4 restored flow^47^; in 540 patients, NET-related biomarkers oscillated diurnally in coincidence with reduced collateral flow and worse outcome; and lower thrombus NET content was associated with better collateral grade in 206 thrombectomy patients^48^. NE-rich thrombi may therefore arise in patients with lower systemic inflammatory burden or better preserved vascular reserve, whose pial collaterals avoid the endothelial dysfunction and rarefaction of chronic vascular inflammation^49–52^. The modest CD61-NE correlation and lower cfDNA in NE-high patients fit this reading but establish neither co-localization nor causality; thrombus NE is thus more plausibly a marker of a collateral-competent vascular phenotype than a causal driver of collateral flow.

### CitH3-rich thrombi and long-term survival

The CitH3-mortality association remained stable across progressive adjustment, and exploratory analyses suggested group separation became more evident beyond the first years, although the limited number of events warrants caution. Because CitH3 was not mirrored by circulating markers, this association is unlikely to be captured by plasma biomarkers alone and may reflect a spatially restricted or biologically controlled immunothrombotic response, potentially influenced by interindividual variability in NET clearance. Consistently, the homozygous Q222R DNase1 variant has been independently associated with increased NET burden and with cardiovascular and all-cause mortality after ST-elevation myocardial infarction^53^. Thrombus maturation also deserves consideration: thrombus age has been reported as the only independent predictor of NET content^54^, older thrombi show greater NETosis extent and poorer outcome^55^, and higher aggregate NET burden has been linked to poorer short-term outcome using outcome-optimized cutoffs^56,57^. Our finding may appear counterintuitive, but CitH3 cannot distinguish extracellular NETs from intracellular citrullination, and median-defined enrichment is not equivalent to a high aggregate burden defined by ROC-derived thresholds. Combining temporal markers of maturation with NETosis markers may disentangle these relationships.

### Implications for biomarker-guided NET-targeted therapies

DNase1 disassembles the extracellular DNA scaffold that stabilizes fibrin-platelet-NET networks, enhancing thrombolysis ex vivo in coronary and cerebral thrombi^23,25,41^ and overcoming resistance specific to platelet-rich thrombi^28^. A NET-rich outer shell can persist after tenecteplase, whereas added DNase1 accelerates thrombolysis, improves cerebral blood flow restoration and reduces infarct volume^58^, which is relevant as tenecteplase increasingly replaces alteplase in LVO stroke. Although not designed to assess stratification, our data imply that trials of NET-targeted therapies should not rely exclusively on circulating biomarkers: thrombus-level profiling could refine stratification, validate intrathrombus thresholds and identify patients likely to benefit from adjunctive DNase1.

### Strengths and limitations

Strengths include the prospective design with systematic sampling and paired plasma-thrombus measurements in one cohort; a whole-cohort strategy avoiding prior subtype assumptions, using quantitative medians to define transparent candidate phenotypes; standardized digital pathology with blinded analysis; progressive adjustment displaying the stability of each association; mediation analysis with bootstrap confidence intervals; incremental discrimination beyond clinical variables; and false discovery rate correction of cross-compartment analyses.

Limitations should be acknowledged. This was a single-center study of moderate size without external validation, limiting generalizability and power for subgroup analyses and infrequent outcomes. The median thresholds are internal, hypothesis-generating cutoffs, not validated decision thresholds. Intrathrombus markers reflect antigen presence rather than functional activity, and CitH3 cannot separate extracellular NETs from intracellular citrullination. Formal inter-observer reproducibility metrics were not pre-specified. Retrieved thrombi may not fully represent in-situ occlusion architecture, particularly the fibrin-platelet outer shell adjacent to the vessel wall, and admission plasma provides only a cross-sectional snapshot. Etiology-specific analyses warrant caution given limited subgroup size, particularly for atherothrombotic stroke. Inclusion was conditional on successful retrieval of analysable thrombus, itself correlated with reperfusion; patients without retrieved thrombus, frequently those without successful recanalization, are not represented, so associations involving reperfusion and collaterals may not generalize to the whole LVO-AIS population. Finally, the observational design precludes causal inference, and these associations should be read as biologically informative phenotypic associations rather than mechanistic demonstrations.

### Conclusions

Intrathrombus NETosis profiling identified prognostically distinct thrombus phenotypes associated with different dimensions of stroke evolution after LVO-AIS thrombectomy. Circulating biomarkers, by contrast, primarily reflected broad structural composition and showed limited correspondence with intrathrombus molecular markers. Local immunothrombotic signatures therefore provide clinically relevant information beyond conventional clinical and procedural variables, and the median-defined approach offers hypothesis-generating reference values for external validation and biomarker-guided studies of NET-targeted strategies.

## Acknowledgments

We thank the patients and their families, the multidisciplinary stroke team for clinical assistance, and the histopathology and laboratory technicians for sample processing.

## Author Contributions

I.L., A.M., P.C., A.G-C. and M.A.M. conceived the project and designed the experiments; B.D-B, P.C, F.O., A.R-G., L.A., A.M-S., M.G-S., F.B., J.A-C., C.G., and Y.S. performed experiments and analyzed data; B.D-B, F.O., A.M-S., M.G-S. and F.B. analyzed data; B.D-B., A.M., P.C., A.G-C., M.A.M. and I.L. prepared the manuscript; and all authors had full access to the data in this study, reviewed drafts of the manuscript, and approved the final version.

## Data Availability Statement

De-identified individual-level data and analysis code (Python) supporting the findings of this study are available from the corresponding author upon reasonable request, subject to institutional approval and data-sharing agreements.

## Novelty and Significance

**What Is Known?**

- Neutrophil extracellular traps (NETs) are constitutive components of large-vessel-occlusion acute ischemic stroke thrombi and contribute to reperfusion resistance, and DNase1-based NET-targeted strategies enhance thrombolysis of retrieved cerebral thrombi ex vivo.
- Circulating NETosis biomarkers (cell-free DNA, MPO-DNA and CitH3-DNA complexes) have been proposed as prognostic tools and stratification markers for NET-targeted therapies in acute ischemic stroke.
- Prior work has typically analyzed plasma or intrathrombus markers in isolation, or has grouped thrombi by predefined etiological or histological subtypes (red versus white, TOAST categories), leaving unresolved whether circulating biomarkers reflect the local immunothrombotic profile of the occluding clot.

**What New Information Does This Article Contribute?**

- Intrathrombus CD61, neutrophil elastase and citrullinated histone H3 define three prognostically distinct thrombus phenotypes that map onto separate dimensions of stroke evolution: functional recovery, collateral circulation and long-term survival, respectively, with each association robust to progressive multivariable adjustment.
- The favourable CD61-associated phenotype is a direct thrombus signal rather than a procedural proxy: counterfactual mediation analysis shows only a minimal indirect effect through complete reperfusion (mTICI 3), with the natural indirect effect close to zero.
- Circulating NETosis biomarkers primarily track macroscopic red-blood-cell/fibrin composition of the retrieved thrombus but do not reflect the local intrathrombus NETosis profile: after FDR correction across 48 plasma-thrombus pairs, no intrathrombus marker correlates positively with its circulating counterpart, indicating that plasma readouts cannot substitute for thrombus-level profiling in biomarker-guided NET-targeted trials.

## Summary

Immunothrombosis is central to large-vessel-occlusion acute ischemic stroke, yet whether circulating NETosis biomarkers reflect the local immunothrombotic composition of the occlusive clot has remained unresolved. In 160 patients treated with mechanical thrombectomy, we integrated quantitative intrathrombus immunohistochemistry for CD61, neutrophil elastase and citrullinated histone H3 with paired plasma NETosis biomarkers and analyzed thrombi as a single quantitative biomarker space rather than through predefined etiological or histological categories. Intrathrombus markers identified three prognostically distinct phenotypes with outcome-specific associations that remained robust across progressive adjustment and were biologically complementary rather than co-linear. Counterfactual mediation showed the CD61-recovery association was a direct thrombus signal rather than an indirect procedural effect. In contrast, circulating NETosis biomarkers tracked the macroscopic RBC-fibrin composition of the retrieved clot but did not reflect the local NETosis profile, and no intrathrombus marker correlated positively with its circulating counterpart after false-discovery-rate correction. These findings support intrathrombus profiling as a biologically informative complement to plasma biomarkers and argue that stratification for NET-targeted therapies should not rely on circulating readouts alone.

